# A nationwide study of 331 rare diseases among 58 million individuals: prevalence, demographics, and COVID-19 outcomes

**DOI:** 10.1101/2023.10.12.23296948

**Authors:** Johan H Thygesen, Huayu Zhang, Hanane Issa, Jinge Wu, Tuankasfee Hama, Ana Caterina Phiho Gomes, Tudor Groza, Sara Khalid, Tom Lumbers, Mevhibe Hocaoglu, Kamlesh Khunti, Rouven Priedon, Amitava Banerjee, Nikolas Pontikos, Chris Tomlinson, Ana Torralbo, Paul Taylor, Cathie Sudlow, Spiros Denaxas, Harry Hemingway, Honghan Wu

## Abstract

**Background:** The Global Burden of Disease study has provided key evidence to inform clinicians, researchers, and policy makers across common diseases, but no similar effort with single study design exists for hundreds of rare diseases. Consequently, many rare conditions lack population-level evidence including prevalence and clinical vulnerability. This has led to the absence of evidence-based care for rare diseases, prominently in the COVID-19 pandemic.

**Method:** This study used electronic health records (EHRs) of more than 58 million people in England, linking nine National Health Service datasets spanning healthcare settings for people alive on Jan 23, 2020. Starting with all rare diseases listed in Orphanet, we quality assured and filtered down to analyse 331 conditions with ICD-10 or SNOMED-CT mappings clinically validated in our dataset. We report 1) population prevalence, clinical and demographic details of rare diseases, and 2) investigate differences in mortality with SARs-CoV-2.

**Findings:** Among 58,162,316 individuals, we identified 894,396 with at least one rare disease. Prevalence data in Orphanet originates from various sources with varying degrees of precision. Here we present reproducible age and gender-adjusted estimates for all 331 rare diseases, including first estimates for 186 (56.2%) without any reported prevalence estimate in Orphanet. We identified 49 rare diseases significantly more frequent in females and 62 in males. Similarly we identified 47 rare diseases more frequent in Asian as compared to White ethnicity and 22 with higher Black to white ratios as compared to similar ratios in population controls. 37 rare diseases were overrepresented in the white population as compared to both Black and Asian ethnicities. In total, 7,965 of 894,396 (0.9%) of rare-disease patients died from COVID-19, as compared to 141,287 of 58,162,316 (0.2%) in the full study population. Eight rare diseases had significantly increased risks for COVID-19-related mortality in fully vaccinated individuals, with bullous pemphigoid (8.07[3.01-21.62]) being worst affected.

**Interpretation:** Our study highlights that National-scale EHRs provide a unique resource to estimate detailed prevalence, clinical and demographic data for rare diseases. Using COVID-19-related mortality analysis, we showed the power of large-scale EHRs in providing insights to inform public health decision-making for these often neglected patient populations.

**Funding:** British Heart Foundation Data Science Centre, led by Health Data Research UK.

**Research in context:** *Evidence before the study:* We have previously published the largest study looking at COVID-19 across rare diseases, but with a sample size of 158 COVID-19 infected rare disease patients and 125 unaffected relatives, from Genomics England, the power of that study was limited. We searched PubMed from database inception to Apr 21, 2023, for publications using the search terms “COVID-19” or “SARS-CoV-2” and “rare disease” or “ORPHANET”, without language restrictions. There are many studies examining the severity of COVID-19 in rare disease patients. However, to date, most studies have focused on a single or a few rare diseases associated with severity of COVID-19, and not taken a comprehensive rare disease wide approach. So far no studies have examined the impact of vaccination on mortality in rare disease patients. Moreover, the sample size used to examine rare diseases is limited in most studies. The largest study we identified included 168,680 individuals but only focused on autoimmune rheumatic disease.

*Added value of this study:* In this study we use national scale EHR data from England to report age and gender adjusted point prevalence for 331 rare diseases, with clinically-validated ICD-10 and/or SNOMED-CT code lists. Among these, 186 (56.2%) diseases did not have existing point prevalence data available in Orphanet. To our knowledge, this is the first time that rare diseases have been examined on a national scale, encompassing a population of over 58 million people. The large sample size provides sufficient statistical power to detect and describe enough carriers of even very rare conditions <1 case per million. Our analysis of COVID-related mortality has demonstrated the clinical relevance of national data for rare diseases. Specifically, we identified eight rare conditions that are associated with a significantly increased risk of mortality from COVID-19, even among fully vaccinated individuals.

*Implication of all the available evidence:* These findings provide robust reproducible prevalence, gender, and ethnicity estimates for disease that may often have been under prioritised, and where such information in most cases was not previously available. Our COVID-19 mortality findings highlight the need for targeted policy and support addressing the high level of vulnerability of these patients to COVID-19.

## Introduction

A key conclusion of the largest study to date examining rare-disease prevalence, by Wakap et al. 2020, was that further registry research and rare-disease codification was needed to improve current prevalence estimates. In their study they examined 6,172 RDs listed in the Orphanet database, an international initiative for rare disease research, and estimated that they affect 3.5 - 5.9% of the world population (equal to 263-446 million or about 1 in 17 people) at any point in time^1^. However, they also pointed out that many prevalence estimates in Orphanet are either unknown, based on a small sample size, or expert opinion, this is likely to be imprecise.

RDs, many of which are debilitating and life-threatening, comprise a spectrum of heterogenous phenotypes sharing an often protracted diagnosis, scarce effective treatments, and broadly low quality of life^2,3^. Their low prevalence renders them inherently hard to study, with little incentive to invest in research on diagnostics and treatment for individual RDs. RD research has long been plagued by numerous challenges including: scarce data and reliance on case and family reports for single diseases, geographical variations, lack of definition consensus, use of heterogeneous epidemiological approaches for case ascertainment, and incomplete codification in healthcare systems^1^. Consequently, many RDs have unknown or imprecise prevalence estimates, preventing data-driven healthcare prioritisation which may be detrimental in events such as the COVID-19 pandemic, and hampering systematic assessment and comparison across healthcare systems.

Previous research has measured the indirect impact of COVID-19 on the life and care of RD patients, highlighting the exacerbated difficulties they face such as healthcare service disruption, difficulty of access due to social distancing and safety concerns, declining mental health due to prolonged diagnosis, treatment interference, and subsequently deteriorating symptoms^4–7^. Direct quantification of the risk of severe COVID-19 outcomes in RD populations remains scant. Rutter et al. examined the increased COVID-19 related death rates among patients with rare autoimmune rheumatic diseases in England using electronic health records (EHRs)^8^. In a population-wide study conducted in Hong Kong, Chung et al. described an increase in risk of COVID-19 related death among hospitalised patients with RD^9^. Similar results were found with respect to the first COVID-19 wave in the UK in our previous study, using data from Genomics England^10^.

Risk factors and clinical characteristics continue to be studied to identify those at high risk of severe COVID-19 outcomes, with a growing emphasis on using whole population data for more precise insights ^11–14^. In many countries including the United Kingdom (UK), high risk groups such as people aged 60 and above, those who are immunocompromised, or with comorbidities associated with increased risk of adverse outcomes have been prioritised for vaccination. However, many patients with RDs were not considered extremely vulnerable due to the lack of reliable data on their risk of adverse COVID-19 outcomes. The UK shielded patient list (those with specified underlying conditions considered to make them clinically extremely vulnerable to the development of severe COVID-19 infections) was created in a data-driven way and refined using a risk prediction tool to consider additional risk groups ^15,16^. While a few rare respiratory conditions (bronchiectasis, cystic fibrosis, alveolitis) and neurological conditions (myasthenia gravis, Huntington’s chorea, multiple sclerosis, motor neurone disease) were included in the model, the majority of RDs were unaccounted. This highlights the need to explore the adverse COVID-19 outcomes among RD patients in a large population cohort, and the necessity for improved and effective codification to capture and easily identify RDs in EHRs. In this context, population scale data is crucial to define and refine RD epidemiological estimates and the risk of severe COVID-19 outcomes in this group. This would be useful not only for risk management and vaccine prioritisation in the COVID-19 pandemic but also for the future health of these patients in general.

Using comprehensive and longitudinal patient health data for more than 58 million people in England, this study aims to 1) report reproducible population prevalence of RDs from EHRs and the demographic characteristics of those individuals affected, 2) determine whether people affected by certain RDs have differences in mortality from COVID-19 infection during a time period spanning multiple variants and vaccine roll out.

## Methods

### Data sources and Study population

This cohort study used nine linked datasets from the English National Health Service (NHS) available within the NHS Digital Trusted Research Environment (TRE), accessed through the CVD-COVID-UK / COVID-IMPACT Consortium. The datasets used were: primary care data from the General Practice Extraction Service Extract for Pandemic Planning and Research (GDPPR)^17^, COVID-19 testing data from the Public Health England Second Generation Surveillance System (SGSS), COVID-19 hospital admission data from Secondary Uses Service (SUS), Hospital Episode Statistics for admitted patient care (HES-APC), adult critical care (HES-CC), and outpatients (HES-OP) and the COVID-19 Hospitalisations in England Surveillance System (CHESS); COVID-19 vaccination status; and mortality information from the Office for National Statistics (ONS) Civil Registration of Deaths. A detailed description of these datasets has been published previously by Wood et al. 2021^11^.

The study start date was 23rd January 2020, the date of the first recorded COVID-19 case in the UK^18^, and the end date was 30th November 2021. We included individuals that were: a) alive at the start of the study, b) registered with a General Practitioner (GP) in England (minimum one patient record in GDPPR) between 1st of January 1990 (first year with good data coverage) and prior to 23rd January 2020, c) associated with a valid person pseudo-identifier enabling data linkage.

### Identification of participants with rare-diseases

Full details on identification of rare-disease participants can be found in supplementary methods, a brief description follows below. Orphanet^19^, an extensive online resource for RDs, was used to identify and define RDs, data was downloaded on the 6^th^ of May 2022. We adopted a stepwise approach to identify RDs that could be accurately mapped to the disease codes available in our data sources. First, we extracted all Orphanet diseases with mappings to ICD-10 or SNOMED-CT, the clinical terminologies used in our data sources.

Second, we applied the disease classification filter available on Orphanet to filter diseases to those included in ‘disorder types’: disease, morphological anomaly, malformation syndrome, or clinical syndrome (n = 5,864). Third, we excluded all diseases for which mapping to ICD10 was classified as “narrow to broad”, since our initial manual validation revealed that most if not all of these codes included a combination of rare and common diseases and hence would result in a high degree of misclassification (note that some of the diseases that were removed also had SNOMED-CT code mappings which were still included).

Fourth, we computed the point prevalence for all the eligible diseases in the TRE (n = 3,817 diseases with at least one affected individual), searching across the linked tables for SNOMED-CT codes in GDPPR, and ICD-10 codes in HES-APC and HES-OP. Fifth, manual curation was performed by a clinician to validate the accuracy of the matching of diseases to SNOMED-CT and ICD-10 codes by considering both the code definitions and the code frequencies from the EHR data. All SNOMED-CT codes were specific to the diseases but some ICD-10 codes were still too broad and introduced misclassification as they matched to both rare and common diseases, these were excluded. This resulted in the inclusion of 331 RDs with highly specific mapping, of which 164 had ICD-10 mappings, 140 had SNOMED-CT mappings and 27 had both (figure S1). Participants were defined as having a rare disease if they had one or more codes of each disease’s ICD-10 or SNOMED-CT codes.

### Estimation of point prevalence of rare diseases in the English population

Point prevalence, estimates per 1,000,000 individuals, was calculated including anyone who were alive and had a RD diagnosis between January 1990 and the study start. Age and gender adjusted prevalence estimates were calculated using the 2021 census data from England as the reference population^20^.

We compared our gender- and age-adjusted estimated prevalence with the point prevalence of RD provided by Orphanet. Giving preference to the Orphanet estimated from the United Kingdom, followed by European estimates (regional or national specific), and worldwide estimates last.

For RDs in our sample identified in 5 or fewer individuals, adjusted prevalence estimates could not be given due to the risk of re-identification; these were compared to Orphanet as having a prevalence of 0.1 per million. To assess the quality and strength of evidence of the Orphanet prevalence data sources, we manually reviewed their study design and methodology.

### COVID-19 phenotyping and vaccine data

To determine the impact of COVID-19 on RD patients we used five previously defined COVID-19 phenotypes^21^ In brief we identified 1) positive SARS-CoV-2 tests, 2) COVID-19 diagnosis recorded in primary care, 3) hospital admissions with a COVID-19 diagnosis, 4) ventilatory support related to COVID-19, and 5) COVID-19 mortality (Supplementary methods for details). Onset of COVID-19 was defined as the date of the earliest COVID-19-event (any of the five phenotypes defined above) for each individual. Date of the outcome was defined as the date of the earliest record of COVID-19 mortality. Time to the event was calculated as the day difference between the date of onset and date of the outcome.

Vaccination status was determined from the COVID-19 vaccination dataset, including all vaccinations administered after Dec 12, 2020 (when the first official dose was administered in England). Patients were classified as fully vaccinated once 14 days had elapsed after their second dose.

### Demographic data

Date of birth, sex and ethnicity were extracted from GDPPR and HES datasets. Age at first RD diagnosis and COVID-19 was calculated as the difference between date of birth and the relevant first RD diagnostic code and/or the first COVID event divided by 365.25. Ethnicity was categorised as per the ONS^22^: Asian or Asian British, Black or Black British, mixed, white, other ethnicities and unknown, mapping ethnicity across primary and secondary care prioritising information from primary care^23^.

Fisher’s exact test was performed to test for difference in sex and ethnicity ratios for the 219 RD with 100 or more affected. Comparing the ratio of patients with a specific RD with the ratio in all unaffected (with regards to the specific condition) in the study population. Ethnicity comparison was done relative to the majority White ethnicity group. Bonferroni correction for multiple testing was applied to give a statistical significance threshold of 0.000228 (0.05 / 219).

### COVID-19-related mortality risk analysis and shielding analysis

We performed a retrospective cohort analysis comparing COVID-19 related mortality in people with a certain RD or a RD category, with matched controls from the general population. Differences in COVID-19 related mortality were addressed with a time-to-event analysis. See supplement methods, for full detail of the following analysis. In brief, The study period for assessing COVID-19 related mortality was set from 2020-09-01 to 2021-11-30.

Cohorts were formed for each RD and RD category, using exact matching on age group, sex, ethnicity and vaccination at the ratio of two controls (not affected by the condition) per individual with a RD. Survival functions were estimated using Kaplan–Meier estimator.

To examine the impact of governmental shielding recommendation on rare-diseases, we compared rare diseases with high risk of COVID-19-related deaths and diseases in the NHS digital shielding list^24^.

## Results

### Identifying rare disease patients

From the 10,563 clinical entities listed on Orphanet (6th of May 2022), we identified and examined 331 disorders where clinical mapping to either SNOMED-CT or ICD-10 was of high specificity (see method section for details) see figure S1.

Our study population comprised 58,162,316 individuals registered with a GP in England and were alive at the onset of the pandemic on January 23, 2020^18^. In this cohort we identified 894,396 individuals with at least one of the 331 rare disorders diagnosed prior to the onset of the pandemic, giving a prevalence of 15,377 per million. In total 7,965 of 894,396 (0.9%) of rare-disease patients died from COVID-19, as compared to 141,287 of 58,162,316 (0.2%) in the full study population cohort from the start of the pandemic until 30th November 2021.

We observed 3,359,077 clinical codes (384 unique codes) associated with the 331 conditions. The majority of these 2,816,568 (83.8%, 216 unique codes) were ICD-10 codes recorded during admitted patient care, 372,620 (11.1%, 167 unique codes) were SNOMED-CT codes from general practitioners and 169,889 (5.1%, 206 unique codes) were ICD-10 codes from hospital outpatient care. Where diagnostic codes were available from both primary and secondary care (ICD-10 and SNOMED), we obtained comparable agreement in prevalence between sources especially for the more frequent conditions such as Myasthenia gravis and Aorta (figure S2).

Numbers of identified patients for each condition and point prevalence adjusted for age and gender for all examined RDs can be found in table 1, and data S1. As expected our age and gender adjusted point prevalence was highly correlated with our raw point prevalence estimates, see figure S2. The most frequent RD in our cohort was *Polymyalgia rheumatica*, a condition that causes pain, stiffness and inflammation in muscles. This condition was predominantly identified in older individuals with 84.5% being above 70. With an adjusted point prevalence of 2831.8 per million (n = 158,648) this condition is more prevalent than what’s considered a RD, according to the UK prevalence definition of 500 or less cases per million^25^.

**Table 1:**
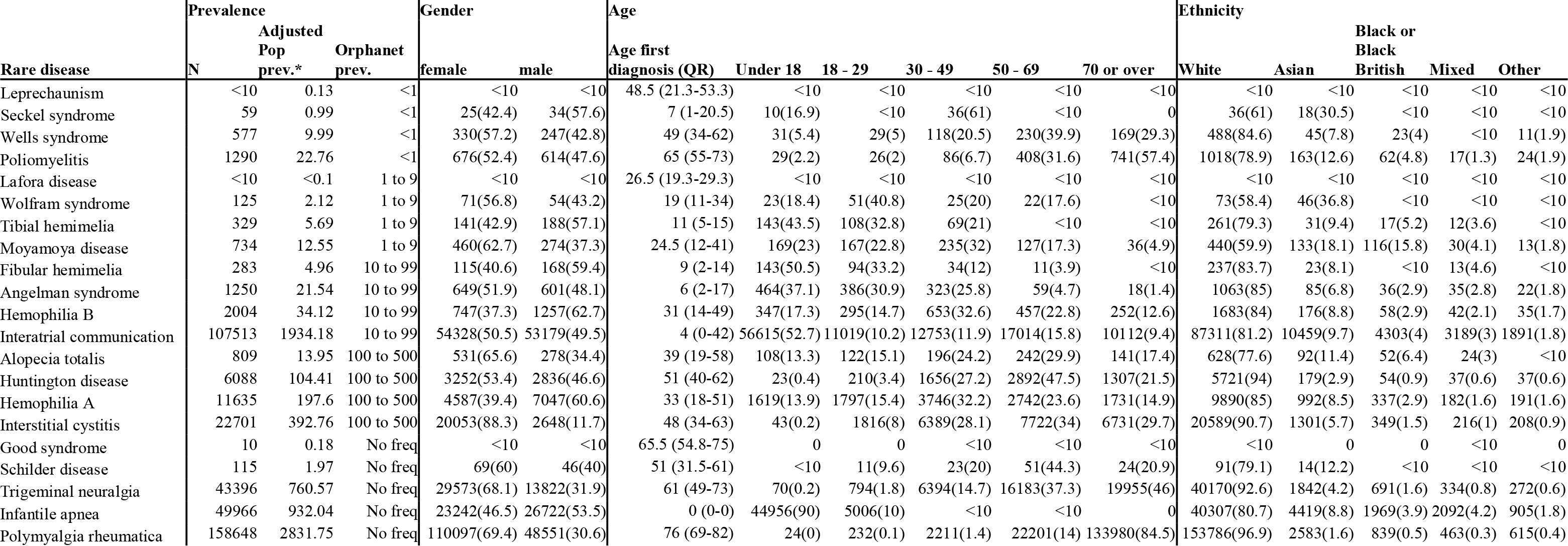
Demographics of 21 selected rare diseases highlighting different prevalence spectra. Table is sorted first by reported Orphanet prevalence per million individuals, category and then study prevalence from low to high. *Adjusted population prevalence is age and gender adjusted prevalence per million individuals. Age is presented as the median age of first diagnosis (25% quantile and 75% quantile) and as numbers and percentages of cases within age groups calculated at study start. Age and sex was available for all diseases. Data for all rare-diseases studied can be seen in data S1 (link to google sheet).

Among all our studied RDs, patients were more often female (55% vs 50.2%), under 18 (25.2% vs 19.9%) or over 70 (28.2% vs 13.1%) as compared to the full study population. There were fewer individuals from the Asian or Asian British (6.7% vs 9.6%) and Black or Black British (2.9% vs 3.9%), communities as compared to the full study population. There were also fewer individuals with unknown ethnicity, which was to be expected from the increased healthcare use and thus registration in the RD group. As expected, RD patients are more likely to be on the shielding list 23.5%, had a SNOMED-CT code indicating shielding, as compared to 7% of the full study population (table 2).

**Table 2:** Demographics table (<u>Link to google sheets version</u>)

| Overall | Study population | All Rare-disease patients | Rare-disease with COVID-19 | Rare-disease with COVID-19 Hospitalisation | Rare-disease with COVID-19 Ventilatory Support | Rare-disease with COVID-19 Deaths |
| --- | --- | --- | --- | --- | --- | --- |
| **n** | 58162316 | 894396 | 128608 | 17750 | 2924 | 7965 |
| **COVID Deaths** | 141287 ( 0.2) | 7965 ( 0.9) | 7965 ( 6.2) | 4641 ( 26.1) | 1322 ( 45.2) | 7965 (100.0) |
| **All Deaths (%)** | 880041 ( 1.5) | 46273 ( 5.2) | 11746 ( 9.1) | 6517 ( 36.7) | 1336 ( 45.7) | 7557 ( 94.9) |
| **Sex (%)** |  |  |  |  |  |  |
| **Female (%)** | 29175170 (50.2) | 492200 (55.0) | 72476 (56.4) | 10377 ( 58.5) | 1464 ( 50.1) | 4425 ( 55.6) |
| **Male (%)** | 28985457 (49.8) | 402171 (45.0) | 56129 (43.6) | 7372 ( 41.5) | 1460 ( 49.9) | 3540 ( 44.4) |
| **Age (%)** |  |  |  |  |  |  |
| Under 18 | 11549034 (19.9) | 225397 (25.2) | 35720 (27.8) | 762 ( 4.3) | 185 ( 6.3) | 17 ( 0.2) |
| 18 - 29 | 8917574 (15.3) | 104538 (11.7) | 18525 (14.4) | 590 ( 3.3) | 110 ( 3.8) | 31 ( 0.4) |
| 30 - 49 | 16158828 (27.8) | 138246 (15.5) | 24234 (18.8) | 1424 ( 8.0) | 397 ( 13.6) | 161 ( 2.0) |
| 50 - 69 | 13894037 (23.9) | 173619 (19.4) | 22172 (17.2) | 3324 ( 18.7) | 1072 ( 36.7) | 1057 ( 13.3) |
| 70 or over | 7642843 (13.1) | 252596 (28.2) | 27957 (21.7) | 11650 ( 65.6) | 1160 ( 39.7) | 6699 ( 84.1) |
| **Ethnic group (%)** |  |  |  |  |  |  |
| White | 46008525 (79.1) | 777730 (87.0) | 111746 (86.9) | 15786 ( 88.9) | 2362 ( 80.8) | 7310 ( 91.8) |
| Asian or Asian British | 5570658 (9.6) | 59717 ( 6.7) | 9584 ( 7.5) | 1081 ( 6.1) | 316 ( 10.8) | 359 ( 4.5) |
| Black or Black British | 2242918 (3.9) | 25935 ( 2.9) | 3172 ( 2.5) | 511 ( 2.9) | 146 ( 5.0) | 159 ( 2.0) |
| Mixed | 1261443 (2.2) | 16652 ( 1.9) | 2337 ( 1.8) | 166 ( 0.9) | 54 ( 1.8) | 47 ( 0.6) |
| Other | 1202022 (2.1) | 10541 ( 1.2) | 1387 ( 1.1) | 180 ( 1.0) | 42 ( 1.4) | 67 ( 0.8) |
| Unknown | 1876750 (3.2) | 3821 ( 0.4) | 373 ( 0.3) | 26 ( 0.1) | <10 | 24 ( 0.3) |
| **Social deprivation fifths (%)** |  |  |  |  |  |  |
| 1 (most deprived) | 11993725 (20.6) | 188047 (21.0) | 29696 (23.1) | 4271 ( 24.1) | 731 ( 25.0) | 1574 ( 19.8) |
| 5 (least deprived) | 11103838 (19.1) | 171851 (19.2) | 23327 (18.1) | 2967 ( 16.7) | 448 ( 15.3) | 1506 ( 18.9) |
| Unknown | 46176 ( 0.1) | 662 ( 0.1) | 60 ( 0.0) | <10 | <10 | <10 |
| **Comorbidities (%)** |  |  |  |  |  |  |
| High risk (Shielding List) | 4096093 ( 7.0) | 210571 (23.5) | 32394 (25.2) | 9578 ( 54.0) | 1614 ( 55.2) | 3397 ( 42.6) |

We compared 219 RD where more than 100 affected individuals were identified, for differences in gender and ethnicity ratios as compared to unaffected in our study population. After Bonferroni correction for multiple testing, we found 111 RD which had significant gender differences. 49 being more prominent in females (replicating known findings for: *Rett syndrome* and *Interstitial cystitis*) and 62 being more prominent in males (examples: *Kennedy’s disease* and *Reactive arthritis*). 100 RD had significant differences between Asian and white ethnicities, with 47 RD being overrepresented in individuals of Asian ethnicity, (*Lamellar ichthyosis* and *Maple urine disease* topped this list). Furthermore, 75 RD had significant differences between Black and white ethnicities, with 22 RD found in higher ratios in the Black community; *Moyamoya disease* and *Quinquaud folliculitis decalvans* had the largest ratios in the Black ethnicities. 37 RD were overrepresented in the white population as compared to both Black and Asian ethnicities (see supplementary data2).

### Comparison of point prevalences with Orphanet

Of the 331 RDs 186 (56.2%) did not have point prevalence data available from Orphanet. Figure 2, compares the gender- and age-adjusted point prevalence observed in our sample with prevalence range estimates from Orphanet, for the 145 (43.8%) diseases with data. This showed that 86 of 145 (59.3%) diseases fall within the same prevalence ranges reported by Orphanet. 25 (17.2%) diseases have higher estimates, with the largest discrepancy observed for interatrial communication: 1,848 per million in our study versus 10-99 per million in Orphanet. 34 (23.4%) of our estimates were lower than Orphanet. For instance, prevalence for chronic actinic dermatitis was 2 per million, while the higher Orphanet estimate is 100-500 per million.

**Figure 2.**
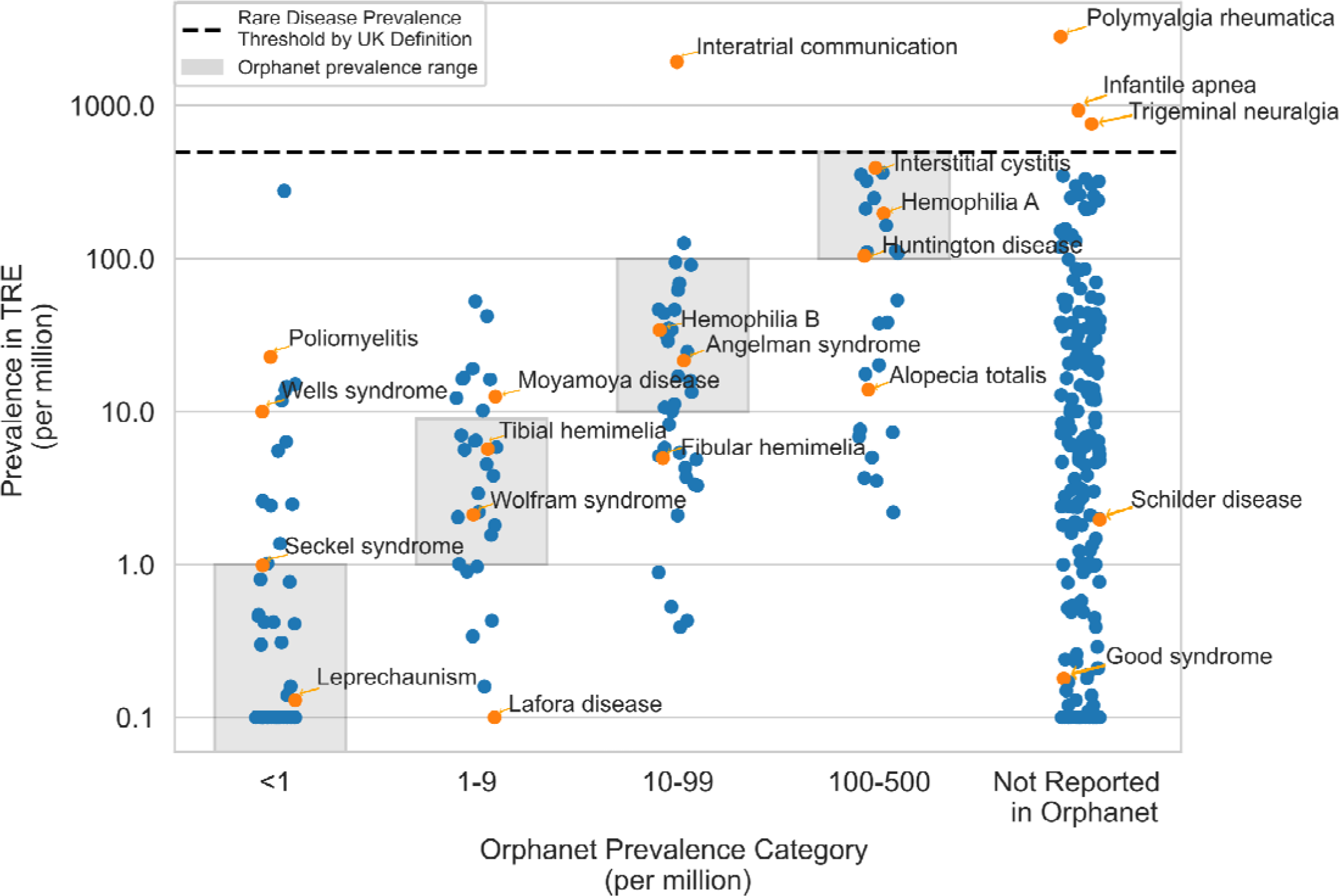
Comparing gender- and age-adjusted rare disease point prevalence estimates from the study data with Orphanet prevalence ranges. The 331 diseases are stratified in groups of relative rarity going from more common to more rare on the x-axis, based on Orphanet estimates. The points indicate our estimated age and gender adjusted point prevalences from population data in England. The labelled diseases are the same 21 selected rare diseases highlighted in table 1. The grey boxes indicate the range of prevalence reported for each disease in Orphanet data. The dashed line represents a prevalence of 1:2000 the upper limit definition for a rare disease suggested by the UK government^25^. COVID19-related mortality in people with rare diseases

To better understand these differences we explored the Orphanet data source of point prevalence estimates for the 145 RDs with data. The data sources were variable and most did not present reproducible method definitions; 63 (43%) had “expert” or “orphanet” as the only source, and could not be verified. Of the 83 with a PubMed Unique Identifier, 23 (28% of 83) were case reports, and 20 (24% of 83) were systematic reviews which might be referring to a case report or an epidemiological study. 28 (34% of 83) referred back to articles where an epidemiological study was conducted, fo 13 (46% of 28) of those no sample size could be found and 3 had no reference data (Table 3).

**Table 3.**
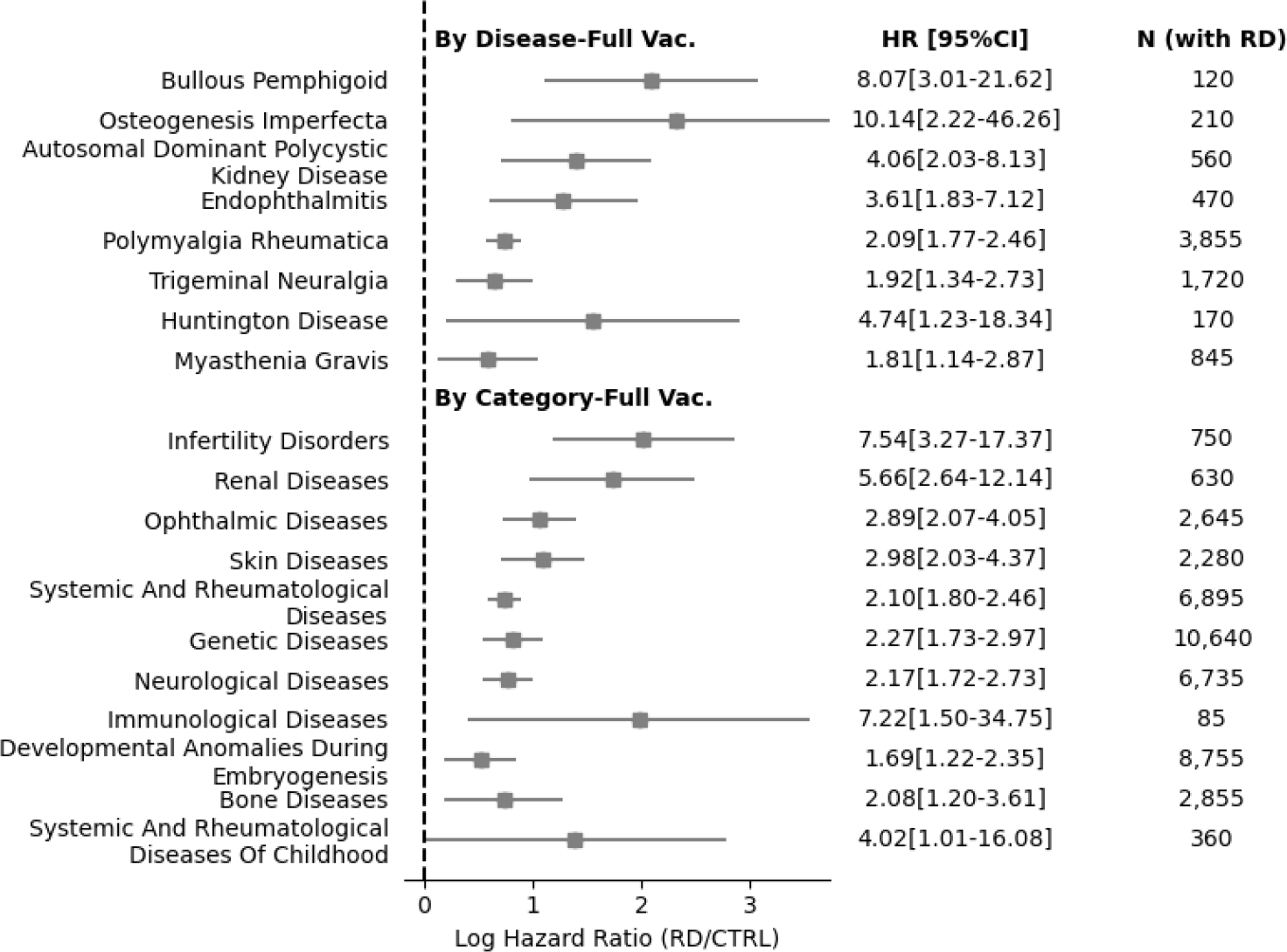
Overview of Orphanet prevalence data sources, used in figure 2 for the 145 (out of 331) rare diseases that had available data. Source types* are not mutually exclusive, some rare diseases have multiple sources listed. Unclear** Little information from these 3 as there is no reference data. Numbers in parentheses are column wise percentages.

### COVID-19 mortality risk

We compared the risk of COVID-19 related death in people with RDs with that of the matched controls from the general population. Of the 331 RDs, compared to matched controls, 8 diseases in vaccinated individuals and 28 in unvaccinated individuals (Figure 3, Supplementary data 3) had significantly increased risk for COVID-19 related death. Of the 25 RD categories (table S1), we observed significantly increased COVID-19 related mortality among vaccinated patients in 11 categories (including 323 diseases of which 179 were included in the analysis) and unvaccinated individuals in 14 categories (including 325 diseases of which 181 were included in the analysis) (Supplementary data 4).

**Figure 3:**
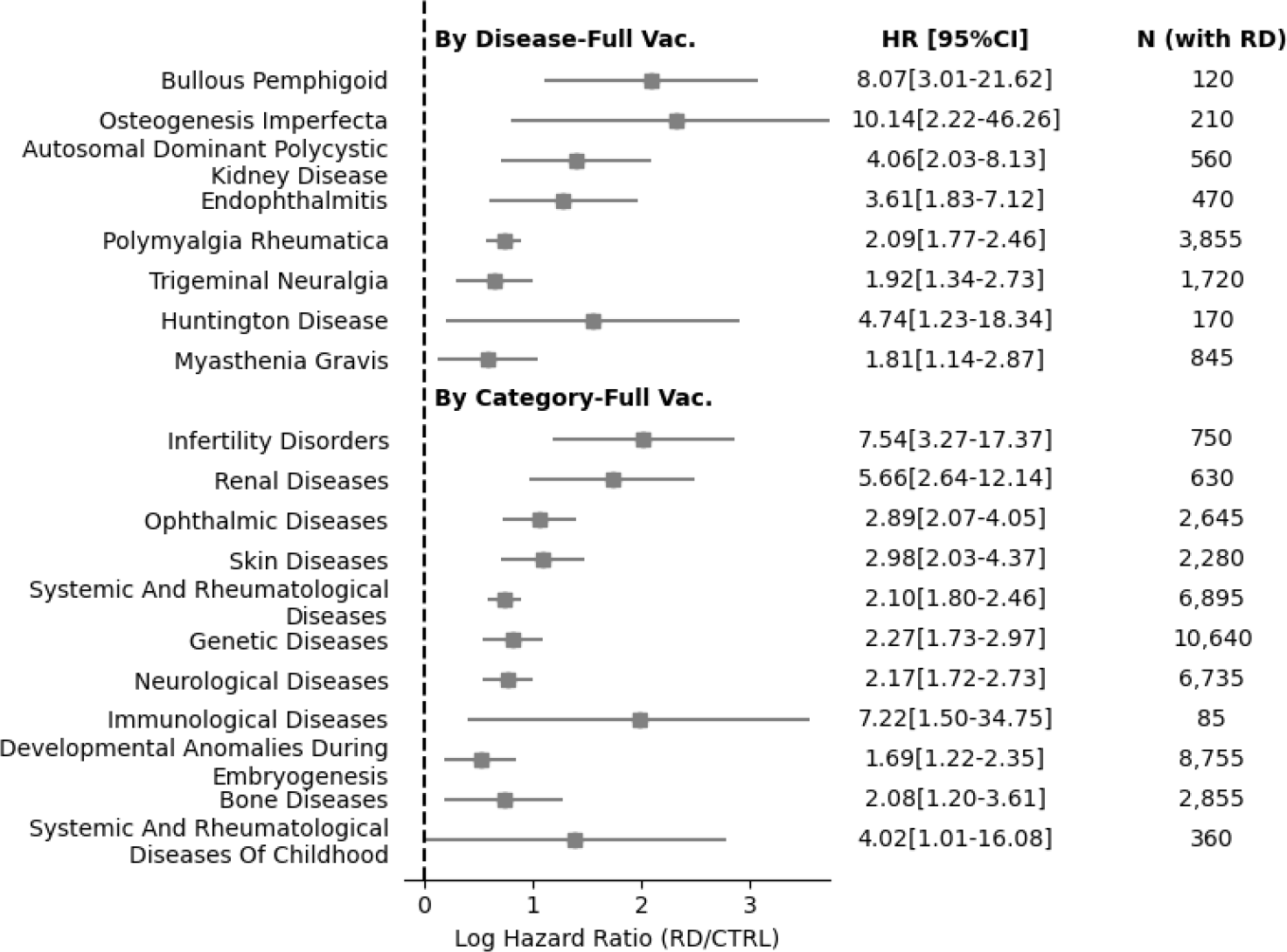
Forest plot showing Log Hazard Ratio of COVID-19-related death for rare diseases and categories with significant increase of risk in fully vaccinated individuals, based on lower-bound of the 95% confidence intervals (95% CI) of the hazard ratio (HR) between individuals with rare diseases (RD) and matched controls (CTRL). HRs and 95% CIs are displayed numerically on the right.

For vaccinated individuals, the three worst affected RDs were bullous pemphigoid (HR [95% CI] = 8.07[3.01-21.62], an autoimmune bullous skin disease), osteogenesis imperfecta (10.14[2.22-46.26], a rare, genetic, primary bone dysplasias), autosomal dominant polycystic kidney disease (4.06[2.03-8.13], a genetic, renal tubular disease). For unvaccinated individuals, the three worst affected RDs were progressive supranuclear palsy (4.11[2.78-6.09], a late-onset neurodegenerative disease), infantile spasms syndrome (16.14[2.02-129.04], a rare epilepsy syndrome) and severe intellectual disability-progressive spastic diplegia syndrome (3.41[1.94-6.02], a genetic, syndromic intellectual disa bility, disorder) (Figure 4). Of the 28 RDs with significantly increased risk of COVID-19 related death in unvaccinated individuals only *Interatrial communication* was included in the list of shield diseases.

**Figure 4:**
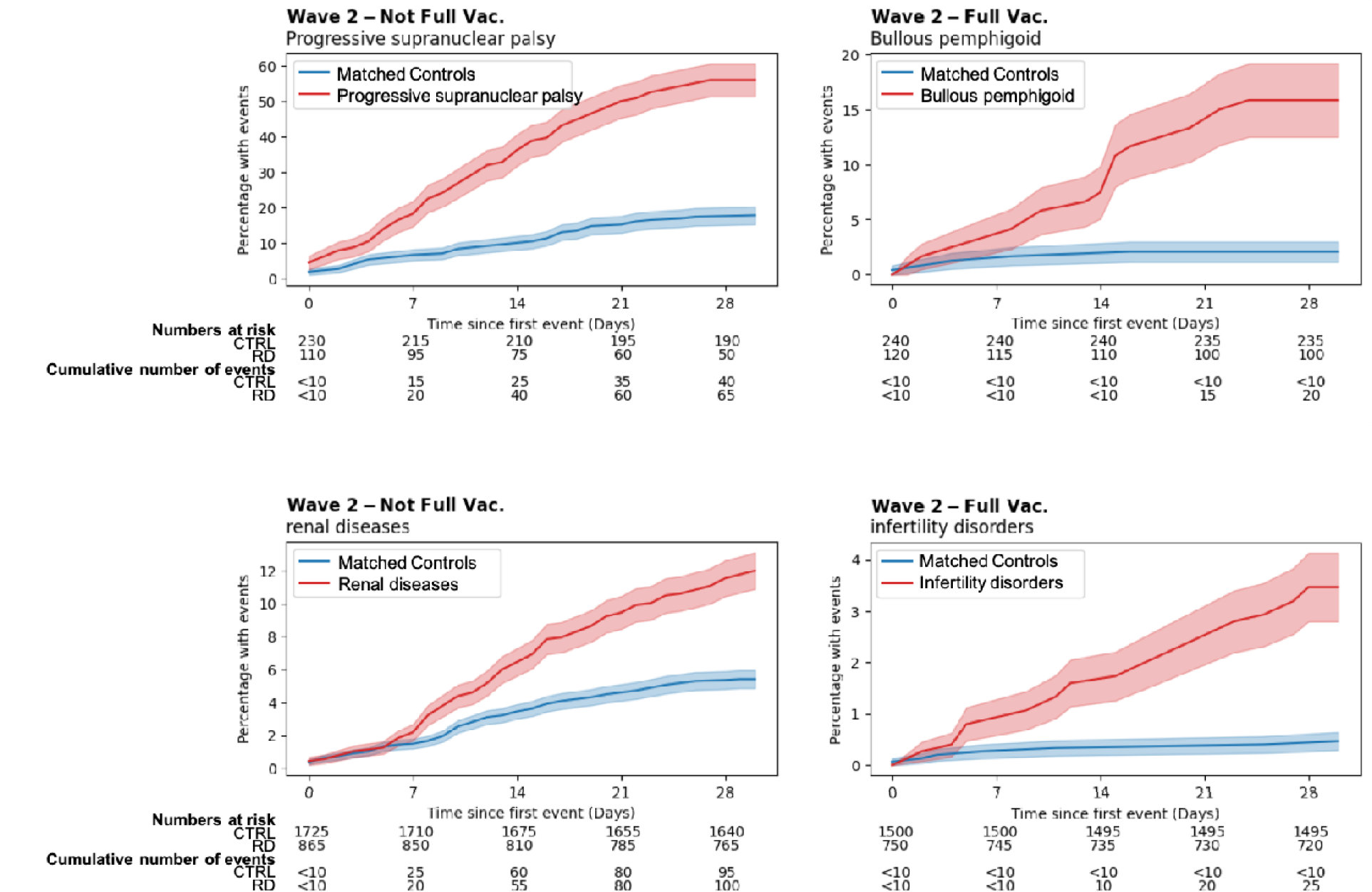
Kaplan-Meier Plot for the most affected rare diseases or categories stratified by if individuals were fully vaccinated, based on lower-bound of the 95% confidence intervals (95% CI) of the hazard ratio (HR) between individuals with rare diseases (RD) and matched controls (CTRL). Events are defined as COVID-related death. Kaplan-Meier Plot for all conditions can be seen in supplementary data 5.

For vaccinated individuals, the three worst affected RD categories were infertility disorders (7.54[3.27-17.37]), renal diseases (5.66[2.64-12.14]), and ophthalmic diseases (2.89[2.07-4.05]). For unvaccinated individuals, the three worst affected RD categories were renal diseases (2.27[1.72-3.00]), infertility diseases (2.21[1.62-3.00]) and ophthalmic diseases (1.82[1.62-2.04]).

Sample size influences power to detect significant HR for COVID-19 mortality in rare disease patients. All significant HR ratios for specific rare diseases were found in disorders which had an adjusted prevalence of more than 17 per million (both for vaccinated and unvaccinated). Significant HR ratios for vaccinated individuals was also only seen in diseases with a later age of onset (median age of first diagnosis > 34 years), as fewer COVID-19 mortalities were observed in younger individuals. However, when considering significant HR estimates based on individuals affected by any rare disease belonging to a certain category, the identified categories with significant HR for COVID-19 mortality in fully vaccinated covered 324 of the 331 rare diseases we examined(Figure 5, panel A).

**Figure 5:**
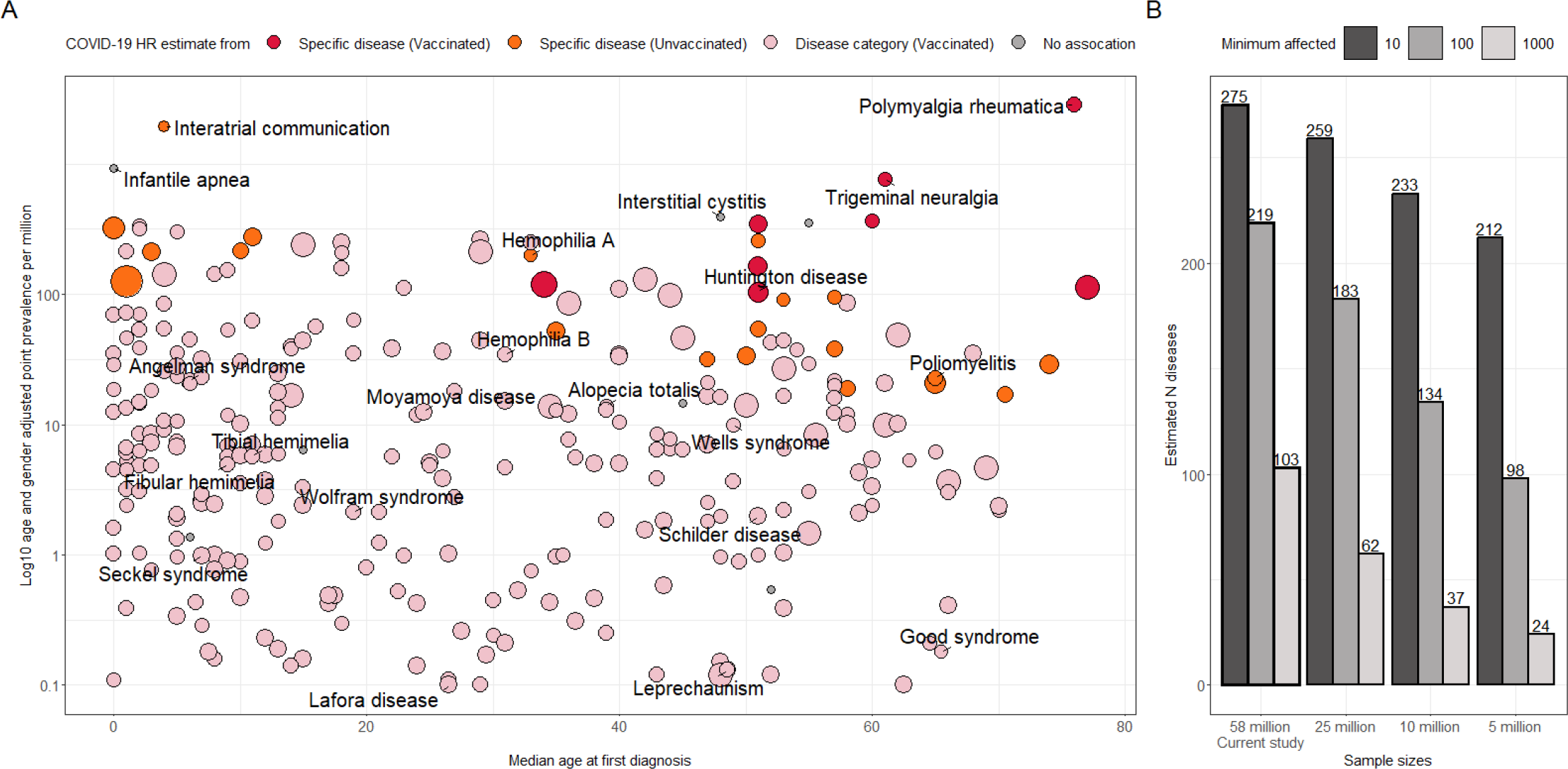
**(A)** Hazard ratio for COVID-19 mortality for 290 Rare diseases in relation to median age at first diagnosis and prevalence. Point sizes are relative to the estimated Hazard Ratio (HR) for COVID-19 mortality. Colours represents the method by which the HR was estimated, and are prioritised in the following order based on what significant HR estimate is available for each disease; (Red, n = 8) estimates from fully vaccinated affected individuals of a specific rare disease, (Orange, n = 20) estimates from none fully vaccinated affected individuals of a specific rare diseases, (Light red, n = 296) estimates based on the highest HR from any of the rare disease category the specific disease belongs to, (dark grey, n = 7) no significant association found for the specific disease or disease category. Age and gender adjusted prevalence rate per million is shown on log10 scale. **(B)** Number of diseases with at least 10, 100 and 1000 identified rare disease patients, observed in our current study sample of ∼58 million, and the estimated numbers of patients if we project our rare disease prevalence into sample sizes of 25, 10, 5 million populations. All current study prevalence estimates are available for all diseases as supplementary data 1.

Lastly, projection of our estimated prevalences into smaller population sizes of 25, 10 and 5 million individuals showcase the need for large scale linked EHR resources in order to identify sufficient rare diseases patients to meaningfully describe these in terms of key demographic strata (Figure 5, panel A).

## Discussion

This study uses linked EHR data for more than 58 million people in England, to provide detailed information for 331 RDs and explore the impact of RDs on COVID-19 mortality. We provide prevalence estimates for 184 RDs which previously did not have such data available from Orphanet. We present, fully reproducible, age and sex adjusted point prevalence estimates for all 331 diseases, including counts of the specific clinical codes observed in our population.

We highlight eight specific RDs where significant increased mortality was observed in fully vaccinated individuals compared to matched individuals from the general population. This is relevant both for countries with high vaccination uptake and those where vaccination rollout is still falling behind. Increased mortality was also seen for 11 out of 25 categories of RD, including RDs where the sizes of individual-disease cohorts did not permit single disease analyses. This paper demonstrates the increased power in RD epidemiology, when linked national data are made available for research.

### Rare-disease selection and case identification

The 331 RDs described in this study is just a fraction of the total number of RDs which have been described in the published literature. Orphanet lists 10,563 clinical entities in their RD database, but many of these (28%) are currently unmapped to clinical codes (ICD-10, SNOMED-CT), complicating case ascertainment from clinical codes. A large proportion (52.5%) of Orphanet listed RD were dropped from our study, either because none of the related codes were observed in our cohort (20.1%), or because the frequency of the mapped clinical codes were excessively common, due to a too broad definition (32.3%). This highlights that even with population wide data, better mappings, to more specific codes, would significantly improve case identification. The more fundamental issue is that classification systems like ICD-10 do not possess the granularity for all RD, or the granularity is not recorded in the clinic. An example of this being that ICD-10 coding is often only recorded to three digits, where the fourth digit level of ICD-10-CM would provide more precision. Orphanet currently only includes mappings to three digit ICD-10 codes in their mappings.

Comprehensive clinical terminologies such as SNOMED-CT or the upcoming ICD-11 may better meet the need for increased specificity, but the use of very specific codes in contemporary health systems remains inconsistent, and systematic efforts will be needed to promote this. For better case identification, previous work has shown the great potential of free-text components of EHRs ^26^, which does however come with its own set of challenges ^27,28^. Phenotypes associated with a disease constitute a computational disease model, is another promising approach to identifying undercoded/undiagnosed RD patients or patients sharing similar vulnerabilities. Both approaches, however, would require more comprehensive and granularEHR data, which currently are not available in national registries, or at least not on a population scale.

### Prevalence comparisons to Orphanet

The European Organization for RDs (Eurordis) and Orphanet have made great progress in assessing the prevalence of RD^32^. However, it is suggested that more reliable data as well as high efficiency with regards to the methodological quality of epidemiological studies is required for the future^32^. Compared to their study, our approach can be generalised for rapid response and continuous monitoring of RD epidemiologic trends, thus filling a gap in the current literature about RD^29^.

The majority of the RD prevalences estimated in our study 184 (55.6%) was not previously known in OrphaData^30^. A significant number of examined Orphanet estimates are not backed by data-driven evidence. Listed sources are not re-assessed, which limits the systematic comparison of estimates due to low inter-study consistency and poor documentation of study methodology. Among diseases with prevalence curated in Orphadata, most of the disease prevalence were consistent (59.6%), although the prevalence of some diseases differed dramatically compared to our study, with both over and under-estimates.

Differences in prevalence estimates may have many causes. Traditionally, prevalence of RD was estimated indirectly from reported incidence^31^ or from meta-analysis of multiple sources of prevalence. Such estimates may be low if there is a poor coverage of incidences. It can also be difficult to estimate the denominator, leading to both under and over-estimates. Some diseases, like interatrial communication, may also be underreported and underestimated due to their asymptomatic nature in early and mild cases. Additionally, the prevalence studies referenced in Orphanet comprise a range of heterogeneous epidemiological approaches, limiting the degree of inter study comparability. For instance, the population from which the published estimate for Chronic Actinic Dermatitis was obtained was restricted to individuals aged 75 and older^32^. Our results highlight the importance of data-driven, reproducible, and age and gender adjusted estimation of RD point prevalence to inform healthcare service planning, clinical trials, pharmacological research, and to allow for precise comparison between studies.

The difference in gender, and ethnicity ratios found for a range of diseases, both replicate known findings and highlight a range of potentially novel insights with regards to differences in disease occurrence between groups, and/or in what may in some cases be disease disparities, indicating inequalities of disease burdens or underdiagnosis in subpopulation groups.

### Strengths and limitations

Statistical power is a major limitation for epidemiologic studies investigating RD. For studies of COVID-19, this issue becomes even more pronounced when the numbers of COVID-19 related mortalities are low, especially in the analyses where individuals were fully vaccinated (visible by larger error bars of the corresponding hazard ratios). Our study utilised large-scale linked EHR data, which greatly improved the statistical power in our analyses.

Our analysis only included demographic covariates for cohort matching. One consideration for this approach is that it is often hard to distinguish whether a non-demographic covariate mediates the effect of RD on risks of COVID-19 related death. Conditioning on such covariates may lead to misleading messages that the people with the RD did not have increased COVID-related mortality. However, for individual RDs, there may be disease-specific covariates that can effectively further stratify people into different risk groups, curation of such information for different diseases is beyond the scope of the current study. We will welcome collaboration of domain-experts to further investigate their diseases of interest.

Despite the comprehensive nature of GDPPR, our cohort included more individuals than the 2021 ONS census population for England (56 489 800)^33^, reflecting the lack of registration data which may allow temporary residents to persist in the dataset, elevating the population denominator and leading to an underestimate of prevalence. Additionally GDPPR contains only a limited subset of SNOMED-CT codes, those which were thought to be related to COVID-19 research^17^, and therefore may underestimate RD prevalence.

It should also be noted that the current analyses focused on the difference of COVID-19 related mortality between people with RDs and the general population. Therefore, the results cannot be seen as an analysis of vaccine effectiveness. Estimation of vaccine effectiveness will be done in a separate study. Further analyses looking at additional outcomes, such as hospitalisation and ventilatory treatment, are also being explored but were out of scope for this study.

## Conclusion

National scale EHRs offer an unprecedented resource to study the epidemiology of RD that may not be possible using other sources. Such basic epidemiological descriptors are crucial for improved planning and prioritisation of RD diagnosis and treatment at national level. This study highlights diseases associated with increasing risk for COVID-19 mortality, which were under-protected by the shielding lists in England at the outbreak of the pandemic. In the current study, we directly and systematically estimated the prevalence of RDs using data-driven methods from large scale linked EHR data. The use of linked primary and secondary data ensured the coverage of RD cases across time and locations. Moreover, our approach is high throughput, allowing the estimation of prevalences of many diseases at the same time, with consistent methodology.

## Data Availability

The authors and colleagues across the CVD-COVID-UK consortium have invested considerable time and energy in developing the data resource described here and are keen to ensure that it is used widely to maximise its value. For inquiries about data access, please see www.healthdatagateway.org/dataset/7e5f0247-f033-4f98-aed3-3d7422b9dc6d or.

## Acknowledgements

This work is carried out with the support of the BHF Data Science Centre led by HDR UK (BHF Grant no. SP/19/3/34678). This work uses data provided by patients and collected by the NHS as part of their care and support. We would like to acknowledge all data providers who make anonymised data available for research.

The views expressed are those of the authors and not necessarily those of the organisations listed. The funders of this work played no role in the collection, analysis, or interpretation of data; in the writing of the report; or in the decision to submit the article for publication.

## Code

The analytical code and protocol are available under an open source licence at the following repository: https://github.com/BHFDSC/CCU019_01

## Data sharing

Dissemination to participants and related patient and public communities: Results will be disseminated through the British Heart Foundation (BHF) Data Science Centre and CVD-COVID-UK webpages on the Health Data Research UK website, BHF communication channels, the BHF Data Science Centre’s lay members panel, and NHS Digital communications channels.

The study team would like to thank the BHF Data Science Centre’s lay members panel for their input and NHS DAE output checkers Lisa Grat, James Walker, Hanna McLean and others.

## Ethical and Regulatory Approvals

The North East-Newcastle and North Tyneside 2 research ethics committee provided ethical approval for the CVD-COVID-UK/COVID-IMPACT research programme (REC No 20/NE/0161). This has been described in detail previously^11^. Data access approval was granted to the CVD-COVID-UK consortium (under project proposal CCU013 High-throughput electronic health record phenotyping approaches) through the NHS Digital online Data Access Request Service^34^ (ref. DARS-NIC-381078-Y9C5K). NHS Digital data have been made available for research under the Control of Patient Information (COPI) notice which mandated the sharing of national electronic health records for COVID-19 research (more info: https://digital.nhs.uk/coronavirus/coronavirus-covid-19-response-information-governance-hub/control-of-patient-information-copi-notice). For further detail see supplementary methods.

## References

1 Nguengang Wakap S, Lambert DM, Olry A, et al. Estimating cumulative point prevalence of rare diseases: analysis of the Orphanet database. Eur J Hum Genet 2020; 28: 165–73.

2 Siegel S, Streetz-van der Werf C, Schott JS, Nolte K, Karges W, Kreitschmann-Andermahr I. Diagnostic delay is associated with psychosocial impairment in acromegaly. Pituitary 2013; 16: 507–14.

3 Green PHR, Stavropoulos SN, Panagi SG, et al. Characteristics of adult celiac disease in the USA: results of a national survey. Am J Gastroenterol 2001; 96: 126–31.

4 Peach E, Rutter M, Lanyon P, et al. Risk of death among people with rare autoimmune diseases compared with the general population in England during the 2020 COVID-19 pandemic. Rheumatology 2021; 60: 1902–9.

5 Lampe C, Dionisi-Vici C, Bellettato CM, et al. The impact of COVID-19 on rare metabolic patients and healthcare providers: results from two MetabERN surveys. Orphanet J Rare Dis 2020; 15: 341.

6 Rare Barometer (EURORDIS survey initiative). How has COVID-19 impacted people with rare diseases? .

7 Chaplin C. Unmasked: an insight into three patients’ rare disease experiences during the COVID-19 pandemic. Orphanet J Rare Dis 2021; 16: 88.

8 Rutter M, Lanyon PC, Grainge MJ, et al. COVID-19 infection, admission and death among people with rare autoimmune rheumatic disease in England: results from the RECORDER project. Rheumatology 2022; 61: 3161–71.

9 Chung CCY, Wong WHS, Chung BHY. Hospital mortality in patients with rare diseases during pandemics: lessons learnt from the COVID-19 and SARS pandemics. Orphanet J Rare Dis 2021; 16: 361.

10 Zhang H, Thygesen JH, Shi T, et al. Increased COVID-19 mortality rate in rare disease patients: a retrospective cohort study in participants of the Genomics England 100,000 Genomes project. Orphanet J Rare Dis 2022; 17: 166.

11 Wood A, Denholm R, Hollings S, et al. Linked electronic health records for research on a nationwide cohort of more than 54 million people in England: data resource. BMJ 2021; 373: n826.

12 BMJ. Reducing barriers to data access for research in the public interest—lessons from covid-19. The BMJ. 2020; published online July 6. https://blogs.bmj.com/bmj/2020/07/06/reducing-barriers-to-data-access-for-research-in-the-public-interest-lessons-from-covid-19/ (accessed Sept 1, 2022).

13 Centers for Disease Control and Prevention. Science Brief: Evidence Used to Update the List of Underlying Medical Conditions Associated with Higher Risk for Severe COVID-19. https://www.cdc.gov/coronavirus/2019-ncov/science/science-briefs/underlying-evidence-table.html.

14 Booth A, Reed AB, Ponzo S, et al. Population risk factors for severe disease and mortality in COVID-19: A global systematic review and meta-analysis. PLoS One 2021; 16: e0247461.

15 Clift AK, Coupland CAC, Keogh RH, et al. Living risk prediction algorithm (QCOVID) for risk of hospital admission and mortality from coronavirus 19 in adults: national derivation and validation cohort study. BMJ 2020; 371: m3731.

16 Reynolds M. How we created the Shielded Patient List. NHS Digital. 2020; published online Nov 17. https://digital.nhs.uk/blog/tech-talk/2020/how-we-created-the-shielded-patient-list (accessed Oct 25, 2022).

17 NHS Digital. General Practice Extraction Service (GPES) data for pandemic planning and research. 2022; published online June 8. https://digital.nhs.uk/coronavirus/gpes-data-for-pandemic-planning-and-research/guide-for-analysts-and-users-of-the-data (accessed Sept 14, 2022).

18 Lillie PJ, Samson A, Li A, et al. Novel coronavirus disease (Covid-19): The first two patients in the UK with person to person transmission. J. Infect. 2020; 80: 578–606.

19 Oprhanet. Orphanet: an online rare disease and orphan drug data base. Oprhanet. http://www.orpha.net (accessed May 5, 2022).

20 Roskams M. Population and household estimates, England and Wales: Census 2021. 2022; published online June 28. https://www.ons.gov.uk/peoplepopulationandcommunity/populationandmigration/populationestimates/datasets/populationandhouseholdestimatesenglandandwalescensus2021 (accessed Feb 2, 2023).

21 Thygesen JH, Tomlinson C, Hollings S, et al. COVID-19 trajectories among 57 million adults in England: a cohort study using electronic health records. Lancet Digit Health 2022; 4: e542–57.

22 Ethnic group, national identity and religion. https://www.ons.gov.uk/methodology/classificationsandstandards/measuringequality/ethnicgroupnationalidentityandreligion (accessed Feb 3, 2023).

23 Pineda-Moncusí M, Allery F, Delmestri A, et al. Digital ethnicity data in population-wide electronic health records in England: a description of completeness, coverage, and granularity of diversity. medRxiv. 2022; published online Nov 11. DOI:10.1101/2022.11.11.22282217.

24 NHS digital; COVID-shielding list documentation - Additions and subtractions. NHS digital (archive). https://webarchive.nationalarchives.gov.uk/ukgwa/20220610000338/ https://digital.nhs.uk/coronavirus/shiel ded-patient-list/methodology/additions-and-subtractions?key= (accessed Feb 2, 2023).

25 Department of Health & Social Care. The UK Rare Diseases Framework. Gov.uk. 2021; published online Jan 9. https://www.gov.uk/government/publications/uk-rare-diseases-framework/the-uk-rare-diseases-framework (accessed Sept 21, 2022).

26 Dong H, Suárez-Paniagua V, Zhang H, Wang M, Whitfield E, Wu H. Rare Disease Identification from Clinical Notes with Ontologies and Weak Supervision. arXiv [cs.CL]. 2021; published online May 5. http://arxiv.org/abs/2105.01995.

27 Groza T, Köhler S, Moldenhauer D, et al. The Human Phenotype Ontology: Semantic Unification of Common and Rare Disease. Am J Hum Genet 2015; 97: 111–24.

28 Foreman J, Brent S, Perrett D, et al. DECIPHER: Supporting the interpretation and sharing of rare disease phenotype-linked variant data to advance diagnosis and research. Hum Mutat 2022; 43: 682–97.

29 Haendel M, Vasilevsky N, Unni D, et al. How many rare diseases are there? Nat Rev Drug Discov 2019; 19: 77–8.

30 Orphanet. Orphadata: Free access data from Orphanet. Orphadata. http://www.orphadata.org (accessed May 5, 2022).

31 Kruger E, McNiven P, Marsden D. Estimating the Prevalence of Rare Diseases: Long-Chain Fatty Acid Oxidation Disorders as an Illustrative Example. Adv Ther 2022; 39: 3361–77.

32 Dawe RS. Chronic actinic dermatitis in the elderly: recognition and treatment. Drugs Aging 2005; 22: 201–7.

33 Population estimates. https://www.ons.gov.uk/peoplepopulationandcommunity/populationandmigration/populationestimates (accessed Feb 14, 2023).

34 NHS Digital. Data Access Request Service (DARS). https://digital.nhs.uk/services/data-access-request-service-dars (accessed May 21, 2021).

