## Supplement for "A nationwide study of 331 rare diseases among 58 million individuals: prevalence, demographics, and COVID-19 outcomes"

**Supplementary methods** **2**
Identification of participants with rare-diseases 2
COVID-19 phenotyping 2
Rare disease COVID-19 analysis 3
COVID-19-related mortality risk analysis 3
Statistical Analysis for COVID-related mortality 3
Comparison of risk of COVID-19-related deaths and high-risk labels 4
**Supplementary Figures** **5**
Supplementary figure 1 - Data processing diagram: 5
Supplementary figure 2 - Comparing prevalence between diagnosis systems: 6
Supplementary figure 3 - Comparing adjusted and raw prevalence estimates: 7
Supplementary figure 4 - Forest plots: 7
**Supplementary data** **10**
Supplementary data 1 - Rare disease demographics: 10
Supplementary data 2 - Gender and ethnicity differences: 10
Supplementary data 3 - COVID-19 mortality for individual rare diseases: 10
Supplementary data 4 - COVID19 mortality analysis for rare disease categories: 10
Supplementary data 5 - All rare diseases - Kaplan-Meier Plot: 10
**Supplementary tables** **12**
Supplementary table 1 - Rare disease information by category: 12
Supplementary table 2 - Overview of Orphanet data sources for prevalence 13
Supplementary table 3 - STROBE statement: 13

### Supplementary methods

#### Identification of participants with rare-diseases

Orphanet[19], an extensive online resource for RDs, was used to identify and define RDs. Its rare disease alignment (from Orphanet code to ICD-10 or SNOMED-CT codes), which is managed by a dedicated information scientist at Orphanet[20,21] was downloaded on the 6th of May 2022. We adopted a stepwise approach to identify RDs that could be accurately mapped to the disease codes available in our data sources. First, we extracted all Orphanet diseases with mappings to ICD-10 or SNOMED-CT (all mapping types included; n = 7,697), the clinical terminologies used in our data sources HES-APC and HES-OP, and GDPPR, respectively. New SNOMED-CT codes were created for RDs in Orphanet terminology mapping exercise.[20] Adaptation of newly defined SNOMED-CT codes in clinical settings can take time, and it is therefore, not surprising that many of these codes were not found used in our EHR data. While many Orphanet listed SNOMED-CT codes are not used in our data sources, those we did find were of high quality with good specificity for identifying RDs according to our manual validation.

Second, to select unique clinical RDs (defined as ‘disorders’ in the Orphanet classification)[2], we applied the disease classification filter available on Orphanet to filter diseases to those included in ‘disorder types’: disease, morphological anomaly, malformation syndrome, or clinical syndrome (n = 5,864). The two largest disorder types removed were *Category* (1,719) and *Clinical subtypes* (769). Third, we excluded all diseases for which mapping to ICD10 was classified as “narrow to broad”, since our initial manual validation revealed that most if not all of these codes included a combination of rare and common diseases and hence would result in a high degree of misclassification (n = 3,384 diseases with ICD-10 mappings were removed, note that some of these diseases also had SNOMED-CT code mappings which were still included).

Fourth, we computed the point prevalence for all the eligible diseases in the TRE (n = 3,817 diseases with at least one affected individual), searching across the linked tables for SNOMED-CT codes in GDPPR, and ICD-10 codes in HES-APC and HES-OP. Fifth, manual curation was performed by a clinician (ACPG) to validate the accuracy of the matching of diseases to SNOMED-CT and ICD-10 codes by considering both the code definitions and the code frequencies from the EHR data. All SNOMED-CT codes were specific to the diseases but some ICD-10 codes were still too broad and introduced misclassification as they matched to both rare and common diseases (e.g., familial Parkinson’s disease mapped to Parkinson’s disease). This resulted in the inclusion of 331 RDs with highly specific mapping, of which 164 had ICD-10 mappings, 140 had SNOMED-CT mappings and 27 had both (figure S1). Participants were defined as having a rare disease if they had one or more codes of each disease’s ICD-10 or SNOMED-CT codes.

#### COVID-19 phenotyping

To determine the impact of COVID-19 on RD patients we used five previously defined COVID-19 phenotypes[23] In brief we identified 1) positive SARS-CoV-2 tests, 2) COVID-19 diagnosis recorded in primary care, 3) hospital admissions with a COVID-19 diagnosis, 4) ventilatory support related to COVID-19, and 5) COVID-19 mortality; including (a) suspected or confirmed COVID-19 diagnosis with ICD-10 term listed anywhere on the death certificate, (b) death within 28 days of the first recorded COVID-19 event, where a COVID-19 diagnosis was not listed anywhere on the death certificate, or (c) a COVID-19 hospital admission with a discharge method or discharge destination denoting death, irrespective of cause and duration after the index event. These COVID-19 phenotypes were identified using clinical codes and data from the following linked tables; SGSS, GDPPR, SUS, HES-APC, HES-CC, CHESS, ONS. Full details of the COVID-19 phenotyping descriptions can be found in the supplementary methods and in the paper by Thygesen et al[23].

### Rare disease COVID-19 analysis

##### COVID-19-related mortality risk analysis

To enable exploration of less prevalent RDs by grouping we adopted the 25 RD categories defined by Orphanet, that allows for diseases to belong to multiple categories. We performed a retrospective cohort analysis comparing COVID-19 related mortality in people with a certain RD or a RD category, with matched controls from the general population. Differences in COVID-19 related mortality were addressed with a time-to-event analysis.

The study period for assessing COVID-19 related mortality was set from 2020-09-01 to 2021-11-30, spanning what is commonly referred to as Wave 2 and the first half of Wave 3 of the pandemic in the UK, driven by the original strain/Alpha variant and Delta variant, respectively[26]. This period reflects a time of the pandemic after which vaccination efforts were rolled out and the COVID-19 testing capacity remained high. A minimum follow-up for each individual was upheld to ensure sufficient time to observe COVID-19 related deaths within 28 days of an infection, in line with the UK Government and Public Health England reported threshold[27].

Individuals whose date of first COVID event fell within the period analysed for COVID-19 mortality were included. Cohorts were formed for each RD, using exact matching on age group, sex, ethnicity and vaccination at the ratio of two controls (not affected by the condition) per individual with a RD.

Analysis by disease categories was done with the identical setting. Analysis by disease categories allows for inclusion of rare-disease individuals affected by a disease too underpowered to be studied on their own. We used the 25 disease categories defined by Orphanet. Cohort formation and matching method was identical with the process for single diseases. An individual could be mapped to multiple disease categories either by being affected by multiple diseases or since one disease can belong to multiple categories.

##### Statistical Analysis for COVID-related mortality

Survival functions were estimated using Kaplan–Meier estimator. Differences in survival distributions between people with RDs and matched controls were tested with the log-rank test. The hazard ratio and confidence interval was estimated using a univariable Cox proportional hazards model, using time of event as the dependent variable and RD status as the independent variable. Proportional hazards assumption was tested using the Schoenfeld residuals[28]. Differences with log-rank test p-value $< 0.05$ and lower-bound of confidence interval of hazard ratios $>1$ were considered significant increases of risk. To determine which rare-disease/category showed the largest difference in mortality to unaffected individuals, a conservative approach of using the lower-bound confidence interval for the hazard ratio for comparison, was adopted. Comparing the lower-bound confidence interval, rather than the main estimate, ensured the top findings did not have large overlapping confidence intervals between affected and unaffected, due to smaller sample sizes.

##### Comparison of risk of COVID-19-related deaths and high-risk labels

Diseases included in the shielding list were obtained from NHS digital[29]. The list of diseases with increased COVID-19 mortality rate in unvaccinated individuals was compared to the shielding list to identify diseases that were missed in the shielding scheme.

### Supplementary Figures

#### Supplementary figure 1 - Data processing diagram:

The diagram depicts the rare-disease inclusion steps. TRE = Trusted Research Environment), GDPPR = General Practice Extraction Service Extract for Pandemic Planning and Research. (editable version can be found here)

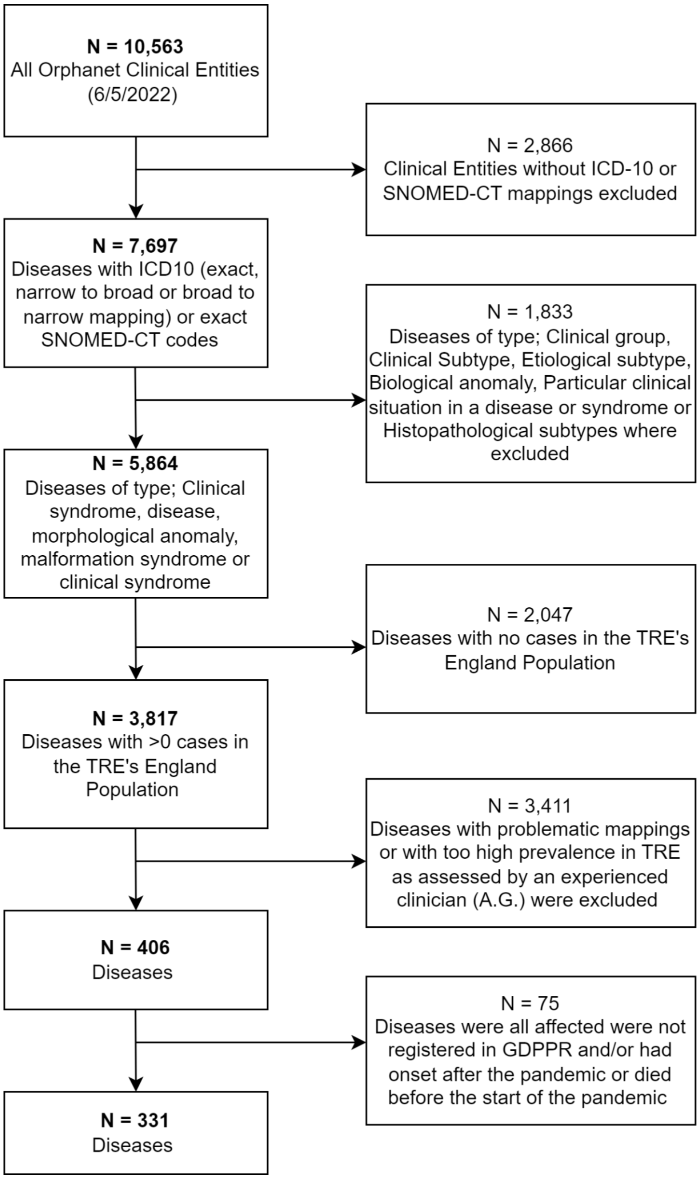

### Supplementary figure 2 - Comparing prevalence between diagnosis systems:

Comparison of prevalence for 27 rare-diseases where affected individuals were identified both from primary care sources (SNOMED-CT) and secondary care data (ICD-10). The left panel shows prevalence per 1,000,000 for all 27 conditions, the dashed line indicating similar prevalence between the sources. The right panel is zoomed in and focused on the less prevalent conditions.

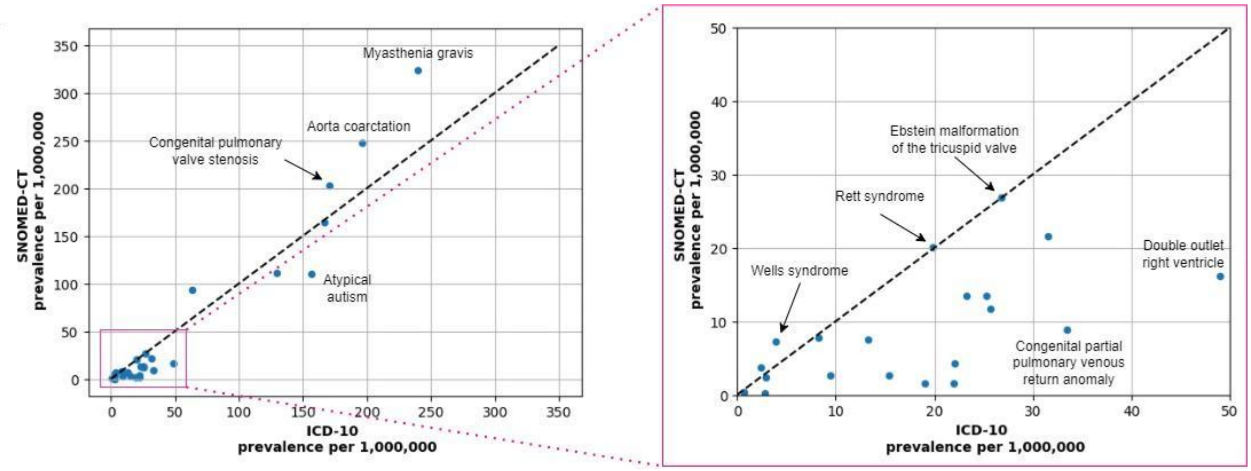

### Supplementary figure 3 - Comparing adjusted and raw prevalence estimates:

Correlation between raw point prevalence per 100.000 on the x-axis, and age and gender adjusted point prevalences per 100.000 (using the English 2021 census data as reference population, see methods for detail). Right plot shows all 331 rare-diseases, left plot is zoomed in showing prevalence smaller than 25 cases per 100.000, for both estimates.

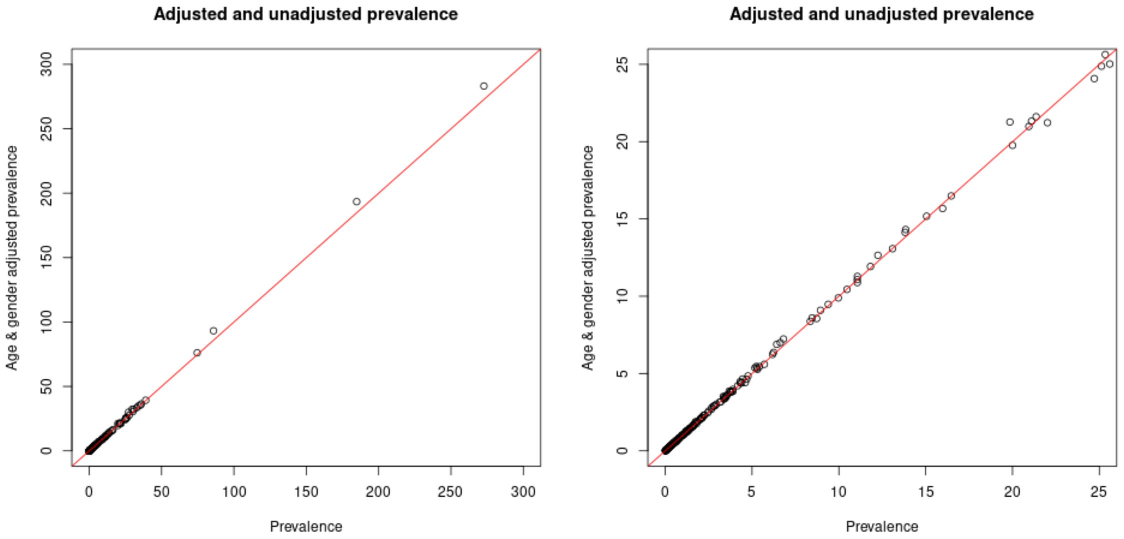

### Supplementary figure 4 - Forest plots:

Forest plot showing Log Hazard Ratio of COVID-19-related death for rare diseases and categories with significant increase of risk in non-fully vaccinated individuals, based on lower-bound of the 95% confidence intervals (95% CI) of the hazard ratio (HR) between individuals with rare diseases (RD) and matched controls (CTRL). HRs and 95% CIs are displayed numerically on the right.

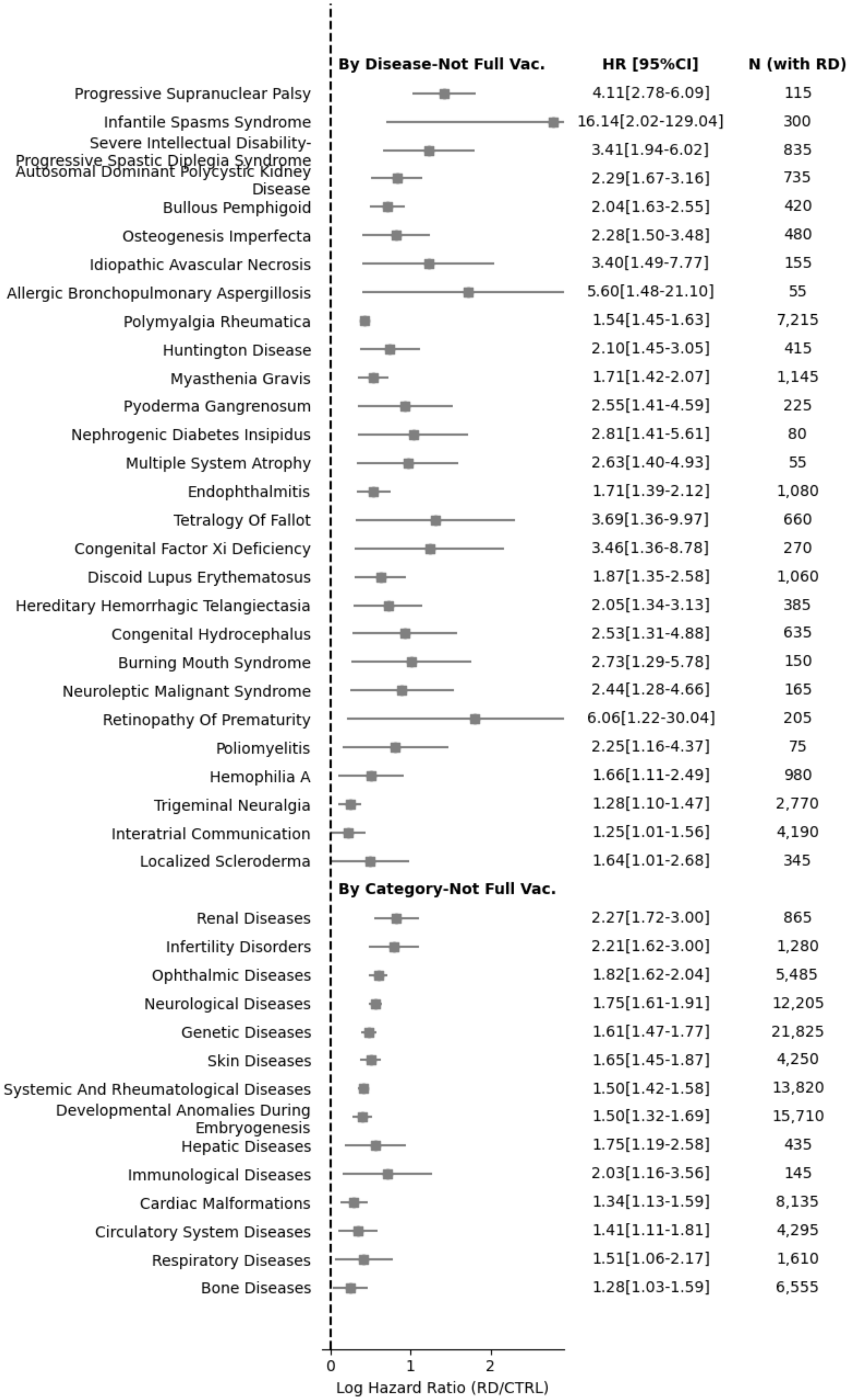
By Disease-Not Full Vac.
HR [95%CI]
N (with RD)
Progressive Supranuclear Palsy 4.11[2.78-6.09] 115
Infantile Spasms Syndrome 16.14[2.02-129.04] 300
Severe Intellectual Disability-Progressive Spastic Diplegia Syndrome 3.41[1.94-6.02] 835
Autosomal Dominant Polycystic Kidney Disease 2.29[1.67-3.16] 735
Bullous Pemphigoid 2.04[1.63-2.55] 420
Osteogenesis Imperfecta 2.28[1.50-3.48] 480
Idiopathic Avascular Necrosis 3.40[1.49-7.77] 155
Allergic Bronchopulmonary Aspergillosis 5.60[1.48-21.10] 55
Polymyalgia Rheumatica 1.54[1.45-1.63] 7,215
Huntington Disease 2.10[1.45-3.05] 415
Myasthenia Gravis 1.71[1.42-2.07] 1,145
Pyoderma Gangrenosum 2.55[1.41-4.59] 225
Nephrogenic Diabetes Insipidus 2.81[1.41-5.61] 80
Multiple System Atrophy 2.63[1.40-4.93] 55
Endophthalmitis 1.71[1.39-2.12] 1,080
Tetralogy Of Fallot 3.69[1.36-9.97] 660
Congenital Factor Xi Deficiency 3.46[1.36-8.78] 270
Discoid Lupus Erythematosus 1.87[1.35-2.58] 1,060
Hereditary Hemorrhagic Telangiectasia 2.05[1.34-3.13] 385
Congenital Hydrocephalus 2.53[1.31-4.88] 635
Burning Mouth Syndrome 2.73[1.29-5.78] 150
Neuroleptic Malignant Syndrome 2.44[1.28-4.66] 165
Retinopathy Of Prematurity 6.06[1.22-30.04] 205
Poliomyelitis 2.25[1.16-4.37] 75
Hemophilia A 1.66[1.11-2.49] 980
Trigeminal Neuralgia 1.28[1.10-1.47] 2,770
Interatrial Communication 1.25[1.01-1.56] 4,190
Localized Scleroderma 1.64[1.01-2.68] 345
By Category-Not Full Vac.
Renal Diseases 2.27[1.72-3.00] 865
Infertility Disorders 2.21[1.62-3.00] 1,280
Ophthalmic Diseases 1.82[1.62-2.04] 5,485
Neurological Diseases 1.75[1.61-1.91] 12,205
Genetic Diseases 1.61[1.47-1.77] 21,825
Skin Diseases 1.65[1.45-1.87] 4,250
Systemic And Rheumatological Diseases 1.50[1.42-1.58] 13,820
Developmental Anomalies During Embryogenesis 1.50[1.32-1.69] 15,710
Hepatic Diseases 1.75[1.19-2.58] 435
Immunological Diseases 2.03[1.16-3.56] 145
Cardiac Malformations 1.34[1.13-1.59] 8,135
Circulatory System Diseases 1.41[1.11-1.81] 4,295
Respiratory Diseases 1.51[1.06-2.17] 1,610
Bone Diseases 1.28[1.03-1.59] 6,555
0
1
2
Log Hazard Ratio (RD/CTRL)

### Supplementary data

#### Supplementary data 1 - Rare disease demographics:

Demographics of all 331 rare diseases. *Adjusted population prevalence is age and gender adjusted prevalence per million individuals. Age is presented as the median age of first diagnosis (25% quantile and 75% quantile) and as numbers and percentages of cases within age groups calculated at study start. Age and sex was available for all diseases.
This table is available as supplementary data 1

#### Supplementary data 2 - Gender and ethnicity differences:

Test statistics for Fishers exact analysis of gender and ethnicity differences of the 219 rare diseases with 100 or more patients identified. Ethnicity analysis compares Asian or Asian British and Black or Black British to the majority White ethnicity.
This table is available as supplementary data 2

#### Supplementary data 3 - COVID-19 mortality for individual rare diseases:

Full results of COVID19 mortality analysis for individual rare diseases. Hazard ratios (HR) were calculated for people with rare diseases to matched controls from the general population. Numbers were rounded to the nearest five and numbers less than ten were suppressed due to data governance requirements.
This table is available as supplementary data 3

#### Supplementary data 4 - COVID19 mortality analysis for rare disease categories:

Full results of COVID19 mortality analysis for rare disease categories. Hazard ratios (HR) were calculated for people with rare diseases to matched controls from the general population. Numbers were rounded to the nearest five and numbers less than ten were suppressed due to data governance requirements.
This table is available as supplementary data 4

#### Supplementary data 5 - All rare diseases - Kaplan-Meier Plot:

Kaplan-Meier Plot for rare diseases or categories stratified by if individuals were fully vaccinated, based on lower-bound of the 95% confidence intervals (95% CI) of the hazard ratio (HR) between individuals with rare diseases (RD) and matched controls (CTRL). Events are defined as COVID-related death.

This figure is available as supplementary data 5

### Supplementary tables

#### Supplementary table 1 - Rare disease information by category:

Percentage of diseases with unknown Orphanet frequency estimates by category. Note that multiple illnesses map to multiple categories, which is why column sums are greater than the total number of examined diseases, see supplementary table 2 for information on which diseases are in each category.

| **Rare disease category** | **331 Rare diseases in category** | **Individuals with at least one disease in the category** | **Mean age of diagnosis** | **Female** | **Male** | **Rare diseases with unknown point prevalence in Orphanet (%)** |
|---|---|---|---|---|---|---|
| Genetic diseases | 202 | 377639 | 23.58 ( 24.18 ) | 181736 (48.1) | 195884 (51.9) | 91 (45) |
| Developmental anomalies during Embryogenesis | 152 | 314917 | 19.03 ( 23.2 ) | 150299 (47.7) | 164608 (52.3) | 94 (61.8) |
| Systemic and rheumatological diseases | 23 | 233241 | 61.7 ( 24.43 ) | 144955 (62.1) | 88285 (37.9) | 16 (69.6) |
| Neurological diseases | 121 | 191779 | 39.95 ( 26.12 ) | 104356 (54.4) | 87417 (45.6) | 47 (38.8) |
| Cardiac malformations | 48 | 175036 | 19.39 ( 24.04 ) | 85419 (48.8) | 89609 (51.2) | 42 (87.5) |
| Ophthalmic diseases | 47 | 111176 | 31.21 ( 29.92 ) | 53427 (48.1) | 57749 (51.9) | 17 (36.2) |
| Bone diseases | 38 | 87126 | 24.77 ( 22.64 ) | 39366 (45.2) | 47759 (54.8) | 25 (65.8) |
| Circulatory system diseases | 22 | 78097 | 17.34 ( 22.54 ) | 36044 (46.2) | 42051 (53.8) | 20 (90.9) |
| Skin diseases | 56 | 60795 | 50.66 ( 22.08 ) | 39792 (65.5) | 21002 (34.5) | 22 (39.3) |
| Respiratory diseases | 8 | 57943 | 7.1 ( 19.5 ) | 27410 (47.3) | 30531 (52.7) | 7 (87.5) |
| Hematological diseases | 9 | 36310 | 33.94 ( 21.95 ) | 20222 (55.7) | 16084 (44.3) | 0 |
| Urogenital diseases | 2 | 27560 | 42 ( 22.46 ) | 20054 (72.8) | 7505 (27.2) | 1 (50) |
| Otorhinolaryngological diseases | 11 | 25748 | 7.37 ( 16.28 ) | 10816 (42) | 14932 (58) | 6 (54.5) |
| Gastroenterological diseases | 4 | 22057 | 52.67 ( 20.06 ) | 11296 (51.2) | 10760 (48.8) | 1 (25) |
| Infertility disorders | 9 | 17265 | 34.88 ( 24.18 ) | 6270 (36.3) | 10994 (63.7) | 5 (55.6) |
| Endocrine diseases | 25 | 14107 | 18.69 ( 19.84 ) | 4333 (30.7) | 9773 (69.3) | 10 (40) |
| Systemic and rheumatological diseases of childhood | 6 | 13764 | 21.67 ( 24.13 ) | 5759 (41.8) | 8004 (58.2) | 3 (50) |
| Odontological diseases | 7 | 13737 | 22.03 ( 15.62 ) | 7885 (57.4) | 5850 (42.6) | 2 (28.6) |
| Renal diseases | 3 | 11569 | 50.59 ( 16.98 ) | 5836 (50.4) | 5733 (49.6) | 0 |
| Inborn errors of metabolism | 18 | 7150 | 21.76 ( 27.15 ) | 3653 (51.1) | 3497 (48.9) | 5 (27.8) |
| Hepatic diseases | 2 | 6368 | 53.66 ( 20.28 ) | 3369 (52.9) | 2999 (47.1) | 0 |
| Cardiac diseases | 4 | 2681 | 24.38 ( 21.47 ) | 1528 (57) | 1153 (43) | 2 (50) |
| Immunological diseases | 14 | 2288 | 43.75 ( 26.64 ) | 1170 (51.1) | 1118 (48.9) | 11 (78.6) |
| Gynecological and obstetric diseases | 4 | 2057 | 16.19 ( 17.83 ) | 1015 (49.3) | 1042 (50.7) | 2 (50) |
| Allergic disease | 3 | 1599 | 60.09 ( 18.74 ) | 797 (49.8) | 802 (50.2) | 3 (100) |

### Supplementary table 2 - Overview of Orphanet data sources for prevalence

| **Orphanet prevalence (per million)** | <1 | 1-9 | 10-99 | 100-500 | All estimates |
|---|---|---|---|---|---|
| **Number of rare disease studies (N=145)** | | | | | |
| **Location** | | | | | |
| UK | 3 | 5 | 10 | 5 | |
| EU | 6 | 18 | 28 | 17 | |
| Worldwide | 43 | 5 | 2 | 4 | |
| **Source type*** | | | | | |
| Review | 7 | 3 | 7 | 3 | |
| Article (observational study, survey) | 6 | 9 | 5 | 5 | |
| Case series report | 16 | 2 | 3 | 2 | |
| Guideline | 0 | 1 | 0 | 1 | |
| European Medicine Agency Report | 0 | 2 | 3 | 7 | |
| Expert | 24 | 10 | 22 | 7 | |
| RCT | 0 | 0 | 0 | 1 | |
| Unclear** | 0 | 3 | 0 | 0 | |

*The categories below are not mutually exclusive, as some rare diseases have multiple sources listed.

**Little information from these 3 as there is no reference data. Otherwise we can merge them with the ‘article’ category.

### Supplementary table 3 - STROBE statement:

STROBE Statement—checklist of items that should be included in reports of observational studies

| | Item No | Recommendation | Page No |
|---|---|---|---|
| **Title and abstract** | 1 | (*a*) Indicate the study's design with a commonly used term in the title or the abstract<br>Retrospective cohort study | 1 |
| | | (*b*) Provide in the abstract an informative and balanced summary of what was done and what was found | 2 |
| **Introduction** | | | |
| Background/rationale | 2 | Explain the scientific background and rationale for the investigation being reported | 3-4 |
| Objectives | 3 | State specific objectives, including any prespecified hypotheses | 6 |
| **Methods** | | | |
| Study design | 4 | Present key elements of study design early in the paper | 5-6, S3-4 |
| Setting | 5 | Describe the setting, locations, and relevant dates, including periods of recruitment, exposure, follow-up, and data collection | 5-6, S2-4 |
| Participants | 6 | (*a*) *Cohort study*—Give the eligibility criteria, and the sources and methods of selection of participants. Describe methods of follow-up<br>*Case-control study*—Give the eligibility criteria, and the sources and methods of case ascertainment and control selection. Give the rationale for the choice of cases and controls<br>*Cross-sectional study*—Give the eligibility criteria, and the sources and methods of selection of participants | 5-6, S2-4 |
| | | (*b*) *Cohort study*—For matched studies, give matching criteria and number of exposed and unexposed<br>*Case-control study*—For matched studies, give matching criteria and the number of controls per case<br><br>Exact match with ratio<br>Matching criteria:<br>Age group<br>Sex<br>Ethnicity<br>Deprivation<br>Smoking<br><br>Comorbidities (Might not be able to match)<br>Medications (Might not be able to match) | 5-6, S2-4 |
| Variables | 7 | Clearly define all outcomes, exposures, predictors, potential confounders, and effect modifiers. Give diagnostic criteria, if | S3 |

| | | | |
|---|---|---|---|
| | | applicable | |
| Data sources/measurement | 8* | For each variable of interest, give sources of data and details of methods of assessment (measurement). Describe comparability of assessment methods if there is more than one group | 5, S2-3 |
| Bias | 9 | Describe any efforts to address potential sources of bias | |
| Study size | 10 | Explain how the study size was arrived at | 5, S2 |
| Quantitative variables | 11 | Explain how quantitative variables were handled in the analyses. If applicable, describe which groupings were chosen and why | S3 |
| Statistical methods | 12 | (*a*) Describe all statistical methods, including those used to control for confounding | 6, S3 |
| | | (*b*) Describe any methods used to examine subgroups and interactions | |
| | | (*c*) Explain how missing data were addressed | |
| | | (*d*) *Cohort study*—If applicable, explain how loss to follow-up was addressed<br>*Case-control study*—If applicable, explain how matching of cases and controls was addressed<br>*Cross-sectional study*—If applicable, describe analytical methods taking account of sampling strategy | S3 |
| | | (*e*) Describe any sensitivity analyses | |

**Results**

| | | | |
|---|---|---|---|
| Participants | 13* | (a) Report numbers of individuals at each stage of study—eg numbers potentially eligible, examined for eligibility, confirmed eligible, included in the study, completing follow-up, and analysed | 8 |
| | | (b) Give reasons for non-participation at each stage | |
| | | (c) Consider use of a flow diagram | |
| Descriptive data | 14* | (a) Give characteristics of study participants (eg demographic, clinical, social) and information on exposures and potential confounders | 8 |
| | | (b) Indicate number of participants with missing data for each variable of interest | |

| | | | |
|---|---|---|---|
| | | (c) *Cohort study*—Summarise follow-up time (eg, average and total amount) | |
| Outcome data | 15* | *Cohort study*—Report numbers of outcome events or summary measures over time | *8-14* |
| | | *Case-control study*—Report numbers in each exposure category, or summary measures of exposure | |
| | | *Cross-sectional study*—Report numbers of outcome events or summary measures | |
| Main results | 16 | (*a*) Give unadjusted estimates and, if applicable, confounder-adjusted estimates and their precision (eg, 95% confidence interval). Make clear which confounders were adjusted for and why they were included | 12 |
| | | (*b*) Report category boundaries when continuous variables were categorized | |
| | | (*c*) If relevant, consider translating estimates of relative risk into absolute risk for a meaningful time period | |
| Other analyses | 17 | Report other analyses done—eg analyses of subgroups and interactions, and sensitivity analyses | |
| **Discussion** | | | |
| Key results | 18 | Summarise key results with reference to study objectives | 15 |
| Limitations | 19 | Discuss limitations of the study, taking into account sources of potential bias or imprecision. Discuss both direction and magnitude of any potential bias | 16 |
| Interpretation | 20 | Give a cautious overall interpretation of results considering objectives, limitations, multiplicity of analyses, results from similar studies, and other relevant evidence | 16 |

| | | | |
|---|---|---|---|
| Generalisability | 21 | Discuss the generalisability (external validity) of the study results | 16 |
| **Other information** | | | |
| Funding | 22 | Give the source of funding and the role of the funders for the present study and, if applicable, for the original study on which the present article is based | 16-17 |

*Give information separately for cases and controls in case-control studies and, if applicable, for exposed and unexposed groups in cohort and cross-sectional studies.

**Note:** An Explanation and Elaboration article discusses each checklist item and gives methodological background and published examples of transparent reporting. The STROBE checklist is best used in conjunction with this article (freely available on the Web sites of PLoS Medicine at http://www.plosmedicine.org/, Annals of Internal Medicine at http://www.annals.org/, and Epidemiology at http://www.epidem.com/). Information on the STROBE Initiative is available at www.strobe-statement.org.
